# “SARS-Cov-2 testing in the United Arab Emirates: Population Attitudes and Beliefs”

**DOI:** 10.1101/2021.02.19.21251841

**Authors:** Latifa Mohammad Baynouna AlKetbi, Nico Nagelkerke, Hanan Abdelbaqi, Fatima ALBlooshi, Mariam AlSaedi, Shamsa Almansoori, Ruqaya AlNuaimi, Amal AlKhoori, Aysha AlAryani, Ahmed Al Jiziri, Naji AlMestika, Mariam AlShamsi, Fatima Kayani, Noura Alblooshi, Shamma AlKhajeh, Ibrahim Al Hammadi, Saeed AlDhahei, Jehan AlFalahi

**Affiliations:** MBBS, Arab Bboard Family Medicine, PhD. Professor, Family Medicine, Abu Dhabi Healthcare Services; Abu Dhabi Healthcare Services; UAEU, Community medicine Department; Abu Dhabi University, Public Health College

**Keywords:** SARS-COV-2, attitudes, believes, public health, Testing

## Abstract

**Objectives:** The United Arab Emirates responded to the SARS-COV-2 pandemic and widely implemented test-and-trace strategy. In this cross-sectional questionnaire-based study 531 subjects presenting for SARS-COV-2 testing were recruited to study population’s beliefs and choices regarding testing and were compared to 156 who never been tested.

**Results:** The community uptake in Abu Dhabi Emirate reached 90% (average of 68% overall). In the great majority it was self-motivated as 6% only had doctor referral. Those who had not taken a test were younger in age (p < 0.001), more likely performing activities such as shopping and eating out (p = 0.001), have a medical illness (p < 0.0001), and working from home (p = 0.005). The tested group reported significantly more agreement with the statement, if someone had negative result no need to stay home or wear mask. In conclusion, SARS-COV-2 testing had extensive coverage and high acceptability in the UAE. Acting on concluded beliefs and attitude are key to ensure the testing coverage efficiency and public empowerment.

## Introduction

The SARS-Cov-2 pandemic has affected the whole world. However, the marked heterogeneity among regions and countries regarding its impact in terms SARS-CoV-2 incidence rate, morbidity and mortality is not clear yet. Clearly, PCR test availability must have played a key role. Countries with test scarcities at the start of the pandemic, such as the UK and The Netherlands, had markedly higher mortality rates than countries such as Germany and South Korea with much better test (and trace) capacities^1^. The United Arab Emirates (UAE) has promptly implemented interventions, such as lockdown, major restructuring of its healthcare services such as allocating few main hospitals in the country for SARS-COV-2 and opening fields hospitals for mild to moderate cases with the objective to mitigate the effects of SARS-COV-2 outbreak that could be one of the strongest in the world and to protect its 10 million population^2^. Although it is difficult to spot the most effective strategy, the UAE has succeeded to maintain SARS-COV-2 mortality level that are among the lowest in the world^3^.

The community-based test, trace and treat strategy could have been essential. Many screening centers are distributed across the UAE that are accessible based on catchment areas. Additionally, home screening for the elderly and families, free laborer clinics operated by dedicated teams linked to occupational health in AbuDhabi healthcare services and regular PCR testing for all healthcare workers were implemented very early in the pandemic. The UAE’s PCR SARS-COV-2 tests over 8 months reached 11.3 per 1000 population in October compared to around 4 per 1000 in May and 4 per 1000 in most in the world^1^.

The community’s awareness, acceptance and cooperation were a key; therefore, this study aims to assess the populations believes and choices toward SARS-COV-2 testing. Additionally, comparing those seeking testing compared to others who did not, could help advise strategies to enhance the acceptability of testing as well as identifying higher risk individuals or groups.

## Methods

This is a cross-sectional study utilising an online survey. The questions in the survey were developed to assess the important domains in the area of testing in the SARS-COV-2 pandemic such as rationale, acceptability and responses to the results and its implications. As this is the first pandemic of this size that mobilized the government and the community, no existing validated questionnaire was found, and questions were developed and selected based on consensus from a group of family physicians with an academic background. The survey was piloted among 12 physicians and administrative staff who provided feedback on content and then piloted on a small number (10) of participants from the public to assess face validity. The survey included demographics, such as age, gender, nationality, marital status, place of residence, education and occupation. Knowledge questions were related to the test’s purpose, target population and accuracy. Attitude questions were trust in the results, willingness to undergo test, the appropriate response to negative or positive test results, test acceptability and intentions to repeat the test. In addition, questions about sources of information about the test, contact history, practice of precautionary measures such as staying at home and social visits, presenting symptoms, if any, and comorbidities were asked. Health lifestyle habits such as smoking, physical activities and diet were inquired about as well.

The sample was collected during the peak of the pandemic, April, May and June 2020. A total of 531 participants who presented to the screening centers to undergo the PCR SARS-COV-2 testing completed the survey. This group was compared with 156 patients who never underwent a SARS-COV-2 PCR. This group was sampled from the Ambulatory Healthcare Services AHS primary care physicians’ panel of patients above 18 years of age.

Survey questions were marginally adjusted to tailor the survey for those who did not get tested. A total of 2900 adult patients’ charts, 100 from each of the 29 centers, were reviewed to collect data on testing but 15 centers patients were called by telephone. Tests that are conducted in any of the screening centers or any government facility will be recorded in SEHA EMR. However, tests done in the private sector are not included. Therefore, out of the 360 called from 15 of the 29 centers, 120 (33%) did not answer or the phone was out of use, 84 (23.3%) underwent testing in a non-government facility and 156 responded.

Regarding required sample size, for a confidence level of 95, a margin of error 5%, 148 participants were required in each group to estimate difference in response between the two groups of 5.5%. The statistical analysis was carried out using SPSS v27. Frequencies, crosstabulation and logistic regression were used.

## Results

The uptake of testing was massive as of the 2900 patients EMR records reviewed; 1995 (68.8%) showed they took the test and 810 (31.7%) did not. Among the 15 centers where patients were called, an additional 84 were found to have undergone test it elsewhere than SEHA, raising their percentage to 71.7%. Among these 15 centers included, the highest coverage was 96% and the lowest was 58% while among the 14 not included, 95% was the highest and 35% the lowest.

The demographics are shown in Table 1. More response came from Al Ain city, Ajman, Sharjah and Dubai compared to other Emirates, constituting 19.8%, 22.6%, 17.1% and 18.3%, respectively. Unscreened subjects were older than the screened subjects, mostly married, more unemployed and females were overrepresented in this group. The screened group were more often UAE nationals, 79.7% compared to 60.3%, and with higher educational levels: 57.9% had completed university degree compared to 38.5% in those who never underwent testing. Unscreened group reported more diabetes (14.1%) and hypertension (9%) compared to the screened subjects (5.1% and 5.5%, respectively).

**Table 1.** Study subjects characteristics.

|  |  | Did not screeningtest | Did the Screening | P value |
| --- | --- | --- | --- | --- |
| Gender | Female | 94 (60.3%) | 259 (49.5%) | 0.019 |
|  | Male | 62 (39.7%) | 264 (50.5%) |  |
| Occupation | Employed | 64(41%) | 310(58.4%) | <0.001 |
|  | Un-Employed | 5(3.2%) | 76(14.3%) |  |
|  | Housewife | 59(37.8%) | 33(6.2%) |  |
|  | Retired | 9(5.8%) | 8(1.5%) |  |
|  | Student | 12(7.7%) | 92(17.3%) |  |
|  | Unskilled | 7(4.5%) | 0(0%) |  |
| Nationality | Non-UAE national | 62(39.7%) | 107(20.3%) | <0.001 |
|  | UAE National | 94(60.3%) | 419(79.7%) |  |
| Educational Status | less than high school | 41(26.3%) | 34(6.5%) | <0.001 |
|  | High School | 55(35.3%) | 186(35.6%) |  |
|  | University or more | 60(38.5%) | 303(57.9%) |  |
| City | Abu Dhabi | 55(35.3%) | 20(3.8%) | <0.001 |
|  | Ajman | 0(0%) | 120(22.6%) |  |
|  | Al Ain | 91(58.3%) | 105(19.8%) |  |
|  | Dubai | 2(1.3%) | 97(18.3%) |  |
|  | Fujairah | 0(0%) | 47(8.9%) |  |
|  | Garbiea | 1(0.6%) | 0(0%) |  |
|  | Ras Al Khaima | 4(2.6%) | 6(1.1%) |  |
|  | Sharjah | 2(1.3%) | 91(17.1%) |  |
|  | Um Al Queen | 0(0%) | 39(7.3%) |  |
|  | Missing | 1(0.6%) | 6(1.1%) |  |
| Marital status | Single | 33(21.2%) | 266(50.8%) | <0.001 |
|  | Married | 120(76.9%) | 243(46.4%) |  |
|  | Divorced | 3(1.9%) | 11(2.1%) |  |
|  | Widowed | 0(0%) | 4(0.8%) |  |
| Age groups | <=30 | 60(41.1%) | 239(58%) | <0.001 |
|  | 31-40 | 23(15.8%) | 104(25.2%) |  |
|  | 41-50 | 24(16.4%) | 49(11.9%) |  |
|  | 51-60 | 19(13%) | 13(3.2%) |  |
|  | 61-70 | 14(9.6%) | (1.7%)7 |  |
|  | >70 | 6(4.1%) | 0(0%) |  |
| Comorbidities | DM | 22(14.1%) | 27(5.1%) | <0.001 |
|  | Asthma | 6(3.8%) | 28(5.3%) |  |
|  | Hypertension | 14(9%) | 29(5.5%) |  |
|  | Respiratory medicine | 0(0%) | 10(1.9%) |  |
|  | Heart Diseases | 1(0.6%) | 6(1.1%) |  |
|  | Renal Diseases | 1(0.6%) | 4(0.8%) |  |
|  | Immune-deficiency diseases or Immune suppressive drugs | 3(2) | 8(1.5) |  |
|  | Vitamin D deficiency | 2(1.3%) | 90(16.9%) |  |
|  | Smoker | 0(0%) | 82(15.4%) |  |
| Total |  | 156 | 531 | 687 |

Table 2 shows the responses to survey questions with the differences between those who underwent the SARS-COV-2 PCR test and those who never did. Both groups agreed with the statement that “testing helps to reduce incidence and mortality through preventing more people getting the infection”, 58.1% and 87.8%, respectively. Moreover, both groups agreed that “the purpose of the test is to detect patients with symptoms” although the tested group was less certain with only 46.5% strongly agreeing compared to 85.3% in the never-tested group. Less trust about the test being able to detect all cases was noted among the never tested group compared to the tested; 78.8% strongly disagreed compared to 29.5%.

**Table 2.**
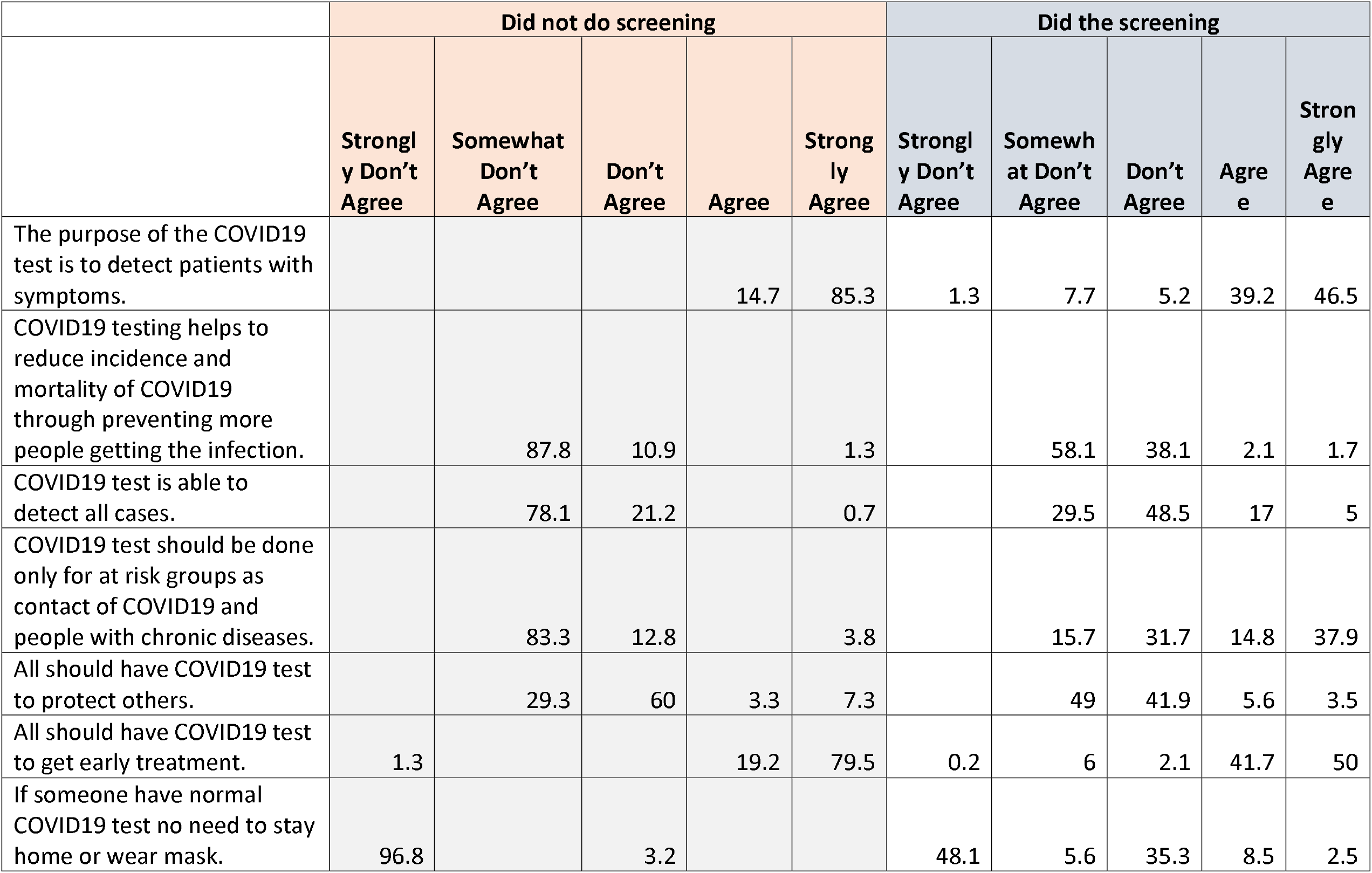
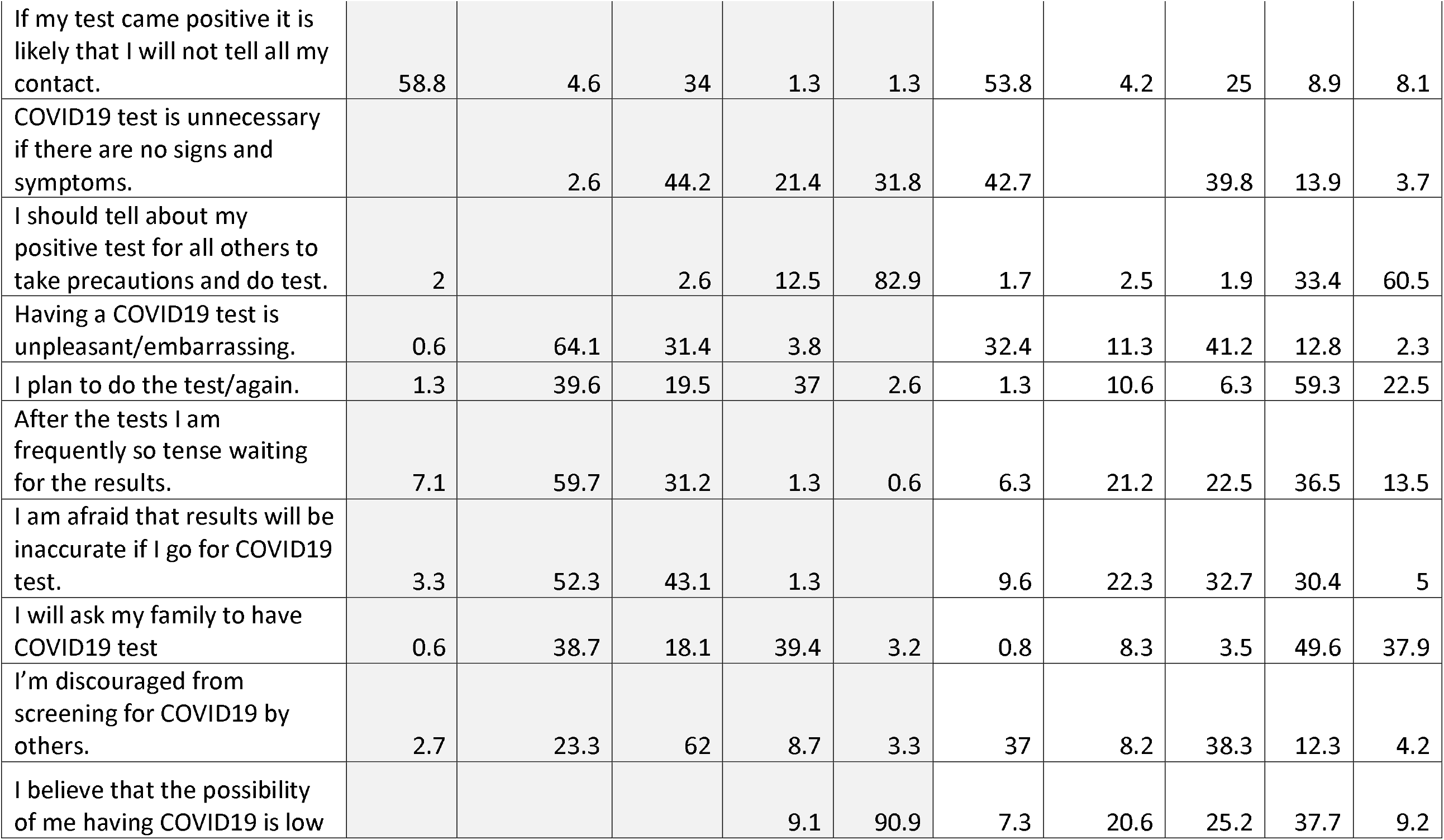
Responses of participants on statements regarding COVID19 testing.

According to Table 3, multivariate logistic regression analysis showed that those who did not undergo the test were significantly engaging in activities such as shopping and eating out more (p=0.001), were younger in age, were more likely of having medical illness (p<0.0001) and more likely working from home (p=0.005). Significant difference in the p value with respect to beliefs between the two groups were observed in responses to questions as follows: “If someone had normal SARS-COV-2 test no need to stay home or wear mask” those who did the screening were significantly more in agreement p value<0.001, 96.8% strongly disagreed among the never-tested compared to 48.1% among the tested group. Surprisingly, those who took the test were more willing to inform any contact if the result is positive, that is 17% compared to 2.6% of those who never tested.

**Table 3.** Significant differences between the studies two groups; those who tested and those who never did.

|  | **B** | **S.E.** | **Sig.** | **OR** | **95% C.I.** |  |
| --- | --- | --- | --- | --- | --- | --- |
| Went out entertaining (garden or eating out) or Shopping in the last 2 weeks | -1.596 | 0.489 | 0.0010 | 0.203 | 0.078 | 0.529 |
| Age | -0.168 | 0.045 | 0.0000 | 0.845 | 0.774 | 0.923 |
| Don't have any medical conditions | -6.14 | 1.505 | 0.0000 | 0.002 | 0 | 0.041 |
| If someone have normal COVID19 test no need to stay home or wear ask. | 2.821 | 0.797 | 0.0000 | 16.79 | 3.518 | 80.121 |
| Having a COVID19 test is unpleasant/embarrassing. | -1.939 | 0.632 | 0.0020 | 0.144 | 0.042 | 0.496 |
| I am afraid that results will be inaccurate if I go for COVID19 test. | 2.259 | 0.639 | 0.0000 | 9.57 | 2.735 | 33.489 |
| I believe that the possibility of me having COVID19 is low | -7.78 | 1.751 | 0.0000 | 0 | 0 | 0.013 |
| Working from Home? | -0.904 | 0.322 | 0.0050 | 0.405 | 0.216 | 0.761 |
| Dependent variable: Doing the COVID19 PCR test |  |  |  |  |  |  |

Additionally, most of those who took the test intend to be tested again or advise their family to be tested, significantly more than the never-tested group. Furthermore, those who had never been tested were significantly more in agreement with “Having a SARS-COV-2 test is unpleasant/embarrassing” statement, 11.3% agreed in the tested group compared to 0.6% in the never tested. Moreover, regarding the statement “I am afraid that results will be inaccurate if I go for a SARS-COV-2 test”, 35.4% of those who did the screening agree compared to 1.3% of those who never tested. Finally, those who never undertook the test were significantly more reassured about not having the infection.

The main reason for not taking the test was “because no one asked us to do it” 70.8%. Other reasons were; because they did not go outside,17.4%, lack of symptoms 6.9%, they did not want to do it, only 2.8% and 1.7% were afraid that staff will infect them. Regarding source of information about testing, those who performed the screening mentioned TV in 22.2% of the responses and 89.1% mentioned social media compared to (1.3%) and (67%) among those who never did the screening respectively. Doctors requested the test for 0.6% in those who did not undergo the test and for 6% in those who did it. Friends and family were the source of advice to do the test in 22.4% and 27.5% of those who took the test and 1.9% and 12.2% of those who never took it respectively. Sponsor was mentioned by 4.3% of those who got tested. Finally, contact with SARS-COV-2 cases was found less likely among those who did not get tested, 92.3% compared to 76.6% among those who did get tested.

Surprisingly, those who never tested were staying home less and participated in social visits more; only 12.5% of them never went to shopping or garden in the last two weeks compared to 55.4% in the tested group (p value =0.001). Moreover, 13.7% had never had visitors or visited others in the last 2 weeks compared to 67% in the tested group.

## Discussion

The very high coverage rate, reaching 90% in some catchment areas and the reponses in this study indicate that the population is aware about the screening, and is motivated towards SARS-COV-2 testing. This could have contributed significantly to the early identification and containment of the outbreak and decreasing mortality and morbidity in the UAE. Nevertheless, there are areas of concern. Abandoning the precautionary restrictions such as wearing mask as a reaction to negative result necessitates health promotion that focuses on such beliefs to ensure no loss of screening gains. These findings relate to the Health Belief Model (HBM) ^4 5^ which posits that messages will achieve optimal behaviour change if they successfully target perceived barriers, benefits, self-efficacy and threat. As the majority of screening was voluntary, the reported high motivation to undergo the test is probably was affected by perceived susceptibility to have SARS-COV-2 with the reported high rate of transmission, perceived severity of the disease and fear of its consequences.

The participants’ responses showed commitment towards the community by participating voluntarily in the screening and the attitude towards positive result where the majority mentioned that they will inform their contacts. Although the minority may not inform contacts probably due to perceived stigma; this can constitute a potential risk of diseases spread and jeopardises screening efforts. Finally, HBM describes the aspect of the self-efficacy which is reported to be high in this study among both groups with high willingness to test if required.

Jones et al. concluded, reflecting on a number of metanalyses, that perceived barriers were the most powerful single predictor of preventive health behaviour across all studies and behaviours, and perceived severity was the least powerful predictor^4 5^. Both perceived susceptibility and perceived benefits were important predictors of protective health behaviour. However, perceived susceptibility was a stronger predictor of preventive health behaviour.

Social media was influential source of information which should be utilised to direct health promotion messages. An area worth directing health promotion towards is the adherence to minimizing social gathering and staying home which was much less practiced by the never-tested group. Lack of perception of susceptibility or threat seems to be the primary factor for not testing regardless of accessibility. It is not poor knowledge, or less exposure to risk but more of a personal choice^6^. This indicates the existence of a risky group which if targeted will decrease SARS-COV-2 transmissions. Nevertheless, the increasing levels of perceived threat of the pandemic and belief in the effectiveness of measures designed to protect against it could further augment their efforts ^6^.

## Conclusion and recommendations

SARS-COV-2 testing had extensive coverage and high acceptability in the UAE. Acting on concluded beliefs and attitude are key to ensure the testing coverage efficiency and public empowerment.

## Limitation

The study was conducted during the first peak of the pandemic. Believes and choices may differ as knowledge accumulate and policy changes. Nevertheless, it is informative to other countries to priorities groups and guide policy.

## Data Availability

Data will be available if requested.

## Declarations

### Ethical approval

This study was approved by the Abu Dhabi COVID19 Research IRB Committee.

### Consent to publish

Abu Dhabi Institutional Research Board approved this publication. All Authors consented.

### Availability of data and materials

Available on request.

### Contributions

Latifa Baynouna AlKetbi conceptualisation data analysis and writing the manuscript, Nico Nagelkerke review of data analysis and manuscript. Hanan Abdelbaqi, Fatima ALBlooshi, Mariam AlSaedi, Shamsa Almansoori, Ruqaya AlNuaimi, Amal AlKhoori, Aysha AlAryani, Ahmed Al Jaziri, Naji AlMestika, Mariam AlShamsi, Fatima Kayani, Noura Alblooshi, Shamma AlKhajeh, Ibrahim Al Hammadi, Jehan AlFalahi,participated in data collection, Saeed AlDhahei data management and manuscript review.

## Acknowledgment

Amna Al Saadi, Mariam Al Kaabi, Muneera Al Blooshi, Raja Farahat, Hodon Saeed, Sameera Omar, Anoud AlShamsi, Mouza Al Dhaheri, Sana Zeinadeen, Mariam Al Kwuiti, Fathya Al Awadhi, Reem Al Falasi.

---

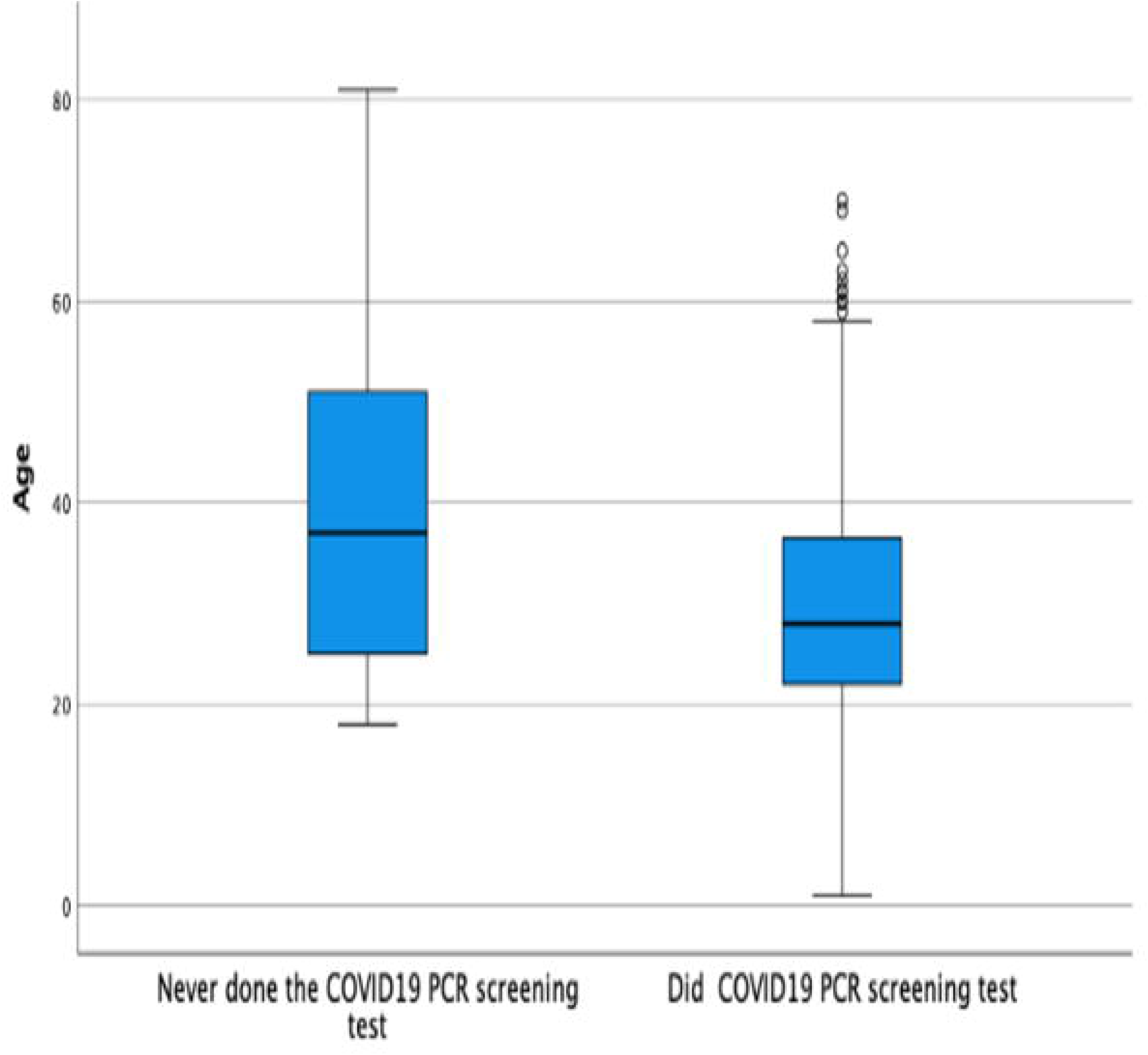

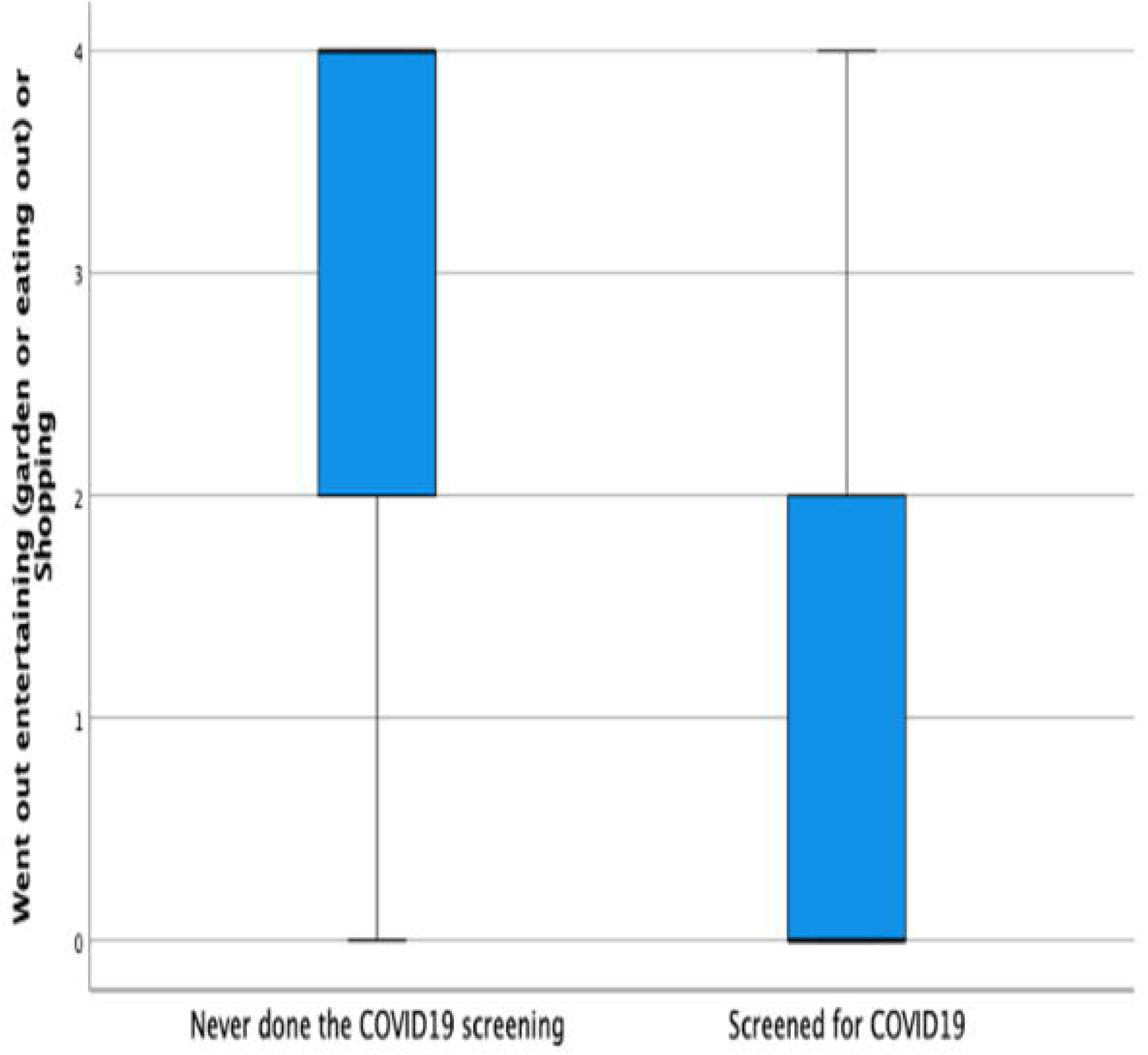

